# Factors associated with the utilisation of mosquito nets amongst rural adults: A cross-sectional study in East Nusa Tenggara Province Indonesia

**DOI:** 10.1101/2023.02.26.23286476

**Authors:** Robertus Dole Guntur, Maria Lobo, Fakir M Amirul Islam

## Abstract

**Backgrounds:** The use of mosquito nets has shown a significant impact on the reduction to malaria burden; however, the utilisation of this kind prevention measures greatly depend on the community behaviour which is limited to be studied in Indonesia. This study explored the factors associated with the use of mosquito nets in the rural of East Nusa Tenggara Province (ENTP) Indonesia.

**Methods:** The community-based cross-sectional study to 1503 rural adults was carried out from October to December 2019. Data on the ownership and the utilisation of mosquito nets were collected using a standardized questionnaire. Logistic regression method was applied to determine the factors associated with the use of mosquito nets.

**Results:** The ownership (utilisation) of any mosquito nets, long-lasting insecticide treated nets (LLINs), and non-LLINs was 95.8% (82.3%), 71.6% (54.8%) and 42.7% (27.6%), respectively. The likelihood of the using of mosquito nets was significantly higher for adults in low malaria endemic settings (MES) than in high MES (adjusted odds ratio [AOR]: 10.1;95% CI, 6.17 to 16.5 for any mosquito nets, AOR: 11.1;95% CI, 7.32 to 16.7 for non-LLINs). The use of LLINs was significantly higher for adults living in Hills areas (AOR: 2.11;95%CI,1.37 to 3.25).

**Conclusion:** The coverage of the ownership nets was not followed by the utilisation of the nets in this province. To progress to malaria elimination in ENTP, the coverage and the utilisation of any mosquito nets should be high. Public health promotion to improve the awareness of the using of these nets should be prioritized, particularly for those living in high MES.

## INTRODUCTIONS

Malaria is most prevalence in low-and middle-income countries ^1^. The World Health Organization (WHO) estimated that the number of malaria cases in 2020 was 241 million ^2^. However, the global trend of malaria deaths decreased about 30% in the last two decades, from 896,000 in 2000 to 627,000 in 2020 ^2^. Based on the WHO report in South East Asia Region, the number of malaria deaths was projected about 8,879 in 2020, of which 83% were from India and 16% were from Indonesia ^2^.

Indonesia which is administratively composed of 514 districts in total has a national commitment to achieve malaria elimination by 2030 ^3^. Sixty-two percent of the total number of districts in the country were classified as malaria elimination zones, while the rest of the districts were classified as low, moderate, and high malaria endemic settings (MES) ^4^. However, this progress shows a significant disparity between Western and Eastern part of the country. Most of districts in the Western part of the country were classified as malaria free zone and low MES. Whilst, most of districts with moderate MES and all districts with high MES were in the Eastern part of the country that includes the East Nusa Tenggara Province (ENTP) ^4^.

ENTP was the third-highest contributor of the malaria burden to the country in 2020 with the total of malaria cases in this province at the end of that year was 15,304 cases ^4^. Under the collaboration with the National Malaria Control Program of Indonesian government, local authority of ENTP has applied various efforts to reduce the burden of malaria in this region. This includes the introduction to apply long lasting insecticide treated net (LLINs) which has been conducted since 2005 ^5^.

The use of LLINs has shown a positive impact on the reduction to malaria burden since 2000 and is considered as the best measure to prevent malaria compared with other malaria prevention approaches ^6^. Therefore, improving the coverage and the utilisation of LLINs is highly recommended by WHO to control malaria vector in malaria endemic countries and the coverage of universal access to LLINs should obtain at least 80% to accelerate malaria elimination effort ^7^. However, in the case the distribution of LLINs was limited, the use of non-LLINS could also be applied as it has also shown a positive impact to reduce the malaria burden ^8,9^

Indonesia applied LLINs as the main vector control intervention in high MES through routine distribution in antenatal care and immunisation activities particularly for pregnant women and children ^3^. The coverage of long-lasting insecticide treated net in high MES has improved significantly from less than 50% in 2007 to 75% in 2017 ^5^. In spite of the fact that the distribution of LLINs increased significantly from 56,337 nets in 2015 to 4,376,636 in 2017 i.e., almost eight times higher, the literature on the coverage and the utilisation of this treated net on the population level was limited in Indonesia ^10^ and the use of this malaria prevention measures greatly depend on human behaviour ^11^. All these yet to be thoroughly investigated in Indonesia.

Some limited study on mosquito nets has been conducted in ENTP [12-16]. A study on village level to explore the behaviour of local community in using mosquito nets indicated that there was an association between family attitude and the use of mosquito nets ^12^. Previous findings related to the current study showed that the knowledge of treated mosquito nets was low [12]-13], and the utilisation of mosquito nets was also low ^15^, as well as the awareness to sleep under mosquito nets varied between ethnicities ^14,16^. However, factors associated with the use of mosquito nets have yet to be investigated in this province. Understanding the contributing factors related to the utilisation of mosquito nets will help the local authority designs an evidence-based approach to boost malaria elimination in this province. This research aimed to fill this gap by exploring the prominent factors associated with the use of mosquito nets to support the national commitment to achieve malaria elimination by 2030.

## METHODS

### Study settings

The current community-based cross-sectional study was conducted in three districts in ENTP from October to December 2019. The eligible participants were 15-85 years of age. Of three districts, firstly, East Sumba district has an estimated number of populations of 258,490 people with 39% had no formal education; secondly, Belu district has an estimated number of populations of 223,176 people with 32.7% had no formal education; and finally, East Manggarai district has an estimated number of population of 287,210 people with 18.4% had no education ^17^

### Study population and sample size calculation

All adults in three selected districts were the target population for this study. In total, the number of respondents enrolled for this study was 1503 adults. This number was obtained after considering of malaria prevalence in ENTP, intra-class correlation of malaria prevention study in Indonesia, design effect and participation rate of participants. The comprehensive calculation of sample size was previously presented elsewhere ^18^.

### Data collection procedure

A data collection tool was adapted from validated questionnaire ^19,20^ with some modifications. Originally the questionnaire was prepared in English. Then, the local language expert and the main author of this study translated the questionnaire into local language (Indonesian). They then combined the translation version of the questionnaire. The combined version of the questionnaire was used as a final tool to collect data. Nine enumerators who had background in nursing school and had working experience in the local public health centres were recruited and trained about the intention of the study, how to approach participants and get their written consent, how to complete the questionnaire. Data collection was supervised strictly by the main author of this study.

### Dependent Variables

The dependent variables of the study were whether participants sleeping under any mosquito net, LLINs, or non-LLINs the night before the survey. These outcomes were obtained based on the response of participants on two questions (did you sleep under mosquito net last night? and what type of mosquito did you use last night?)

### Independent Variables

The independent variables of the study were gender, age group, education level, occupation, family size, socio-economic status (SES), the nearest health facilities, the distance to the nearest health facilities, household income in relation to the provincial minimum wages, household income in relation to Indonesia poverty line and geographical condition.

### Data Analysis

Socio-demographic factors, including age, gender and education of participants were reported by descriptive statistics. The percentage of participants having the possession and utilisation of mosquito nets was computed with its 95% confidence interval (CI). To investigate the potential factors affecting the utilisation of mosquito net, a univariate and multivariate binary logistic regression analysis was applied. Adjusted odds ratio with 95% CI and p value less than 5% was used to affirm the significance of each variable.

### Ethics Statement

This research was approved by Human Ethics Committee of the Swinburne University of Technology (reference number 20191428-1490) and the Indonesian Ministry of Health (reference letter: 164 LB.02.01/2/KE.418/2019). Informed consent was submitted by all participants when they were enrolled.

## RESULTS

The distribution of participants having the ownership of mosquito nets based on the socio-demographic and environmental background is presented in Table 1. People with ownership of mosquito nets were younger, had better education level, Entrepreneurs or government or non-government employees, better SES, from larger family size, and from high or low malaria endemic settings.

**Table 1.**
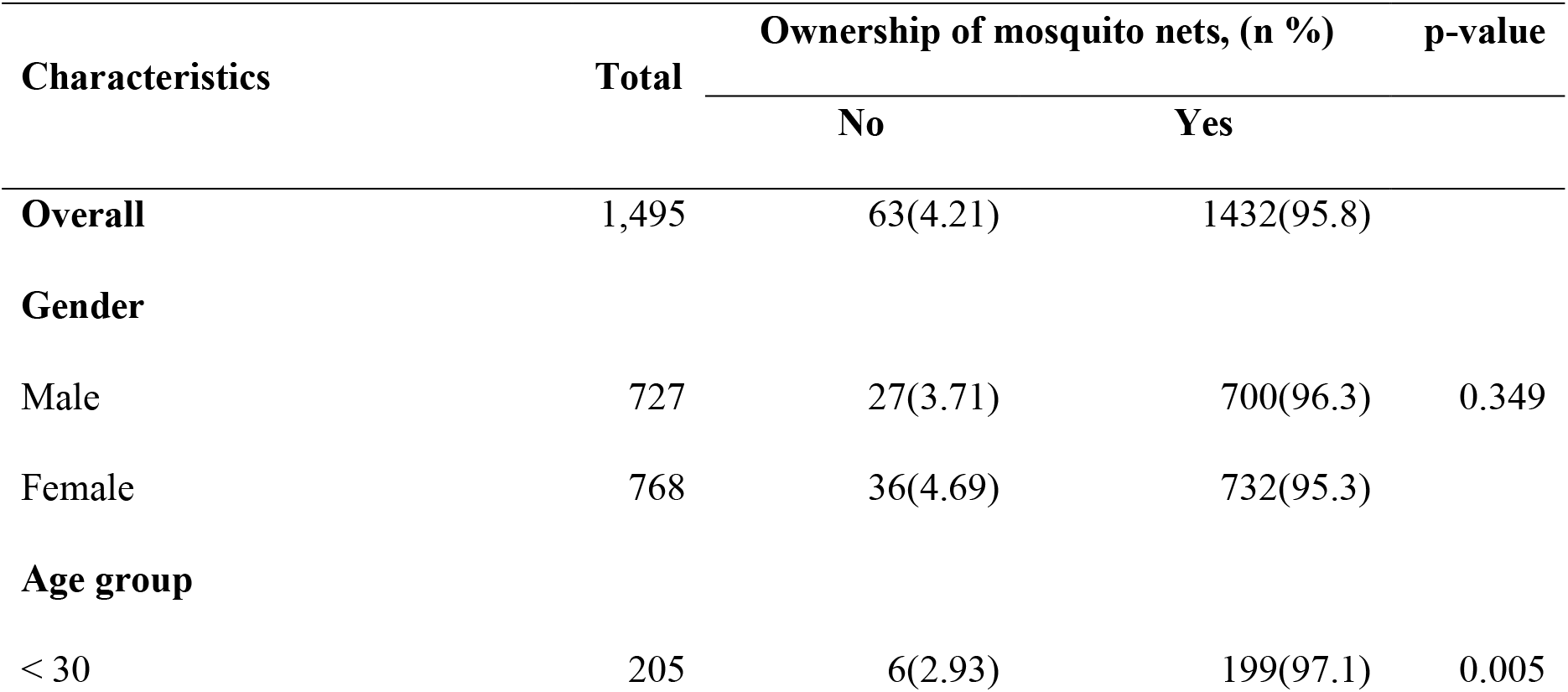
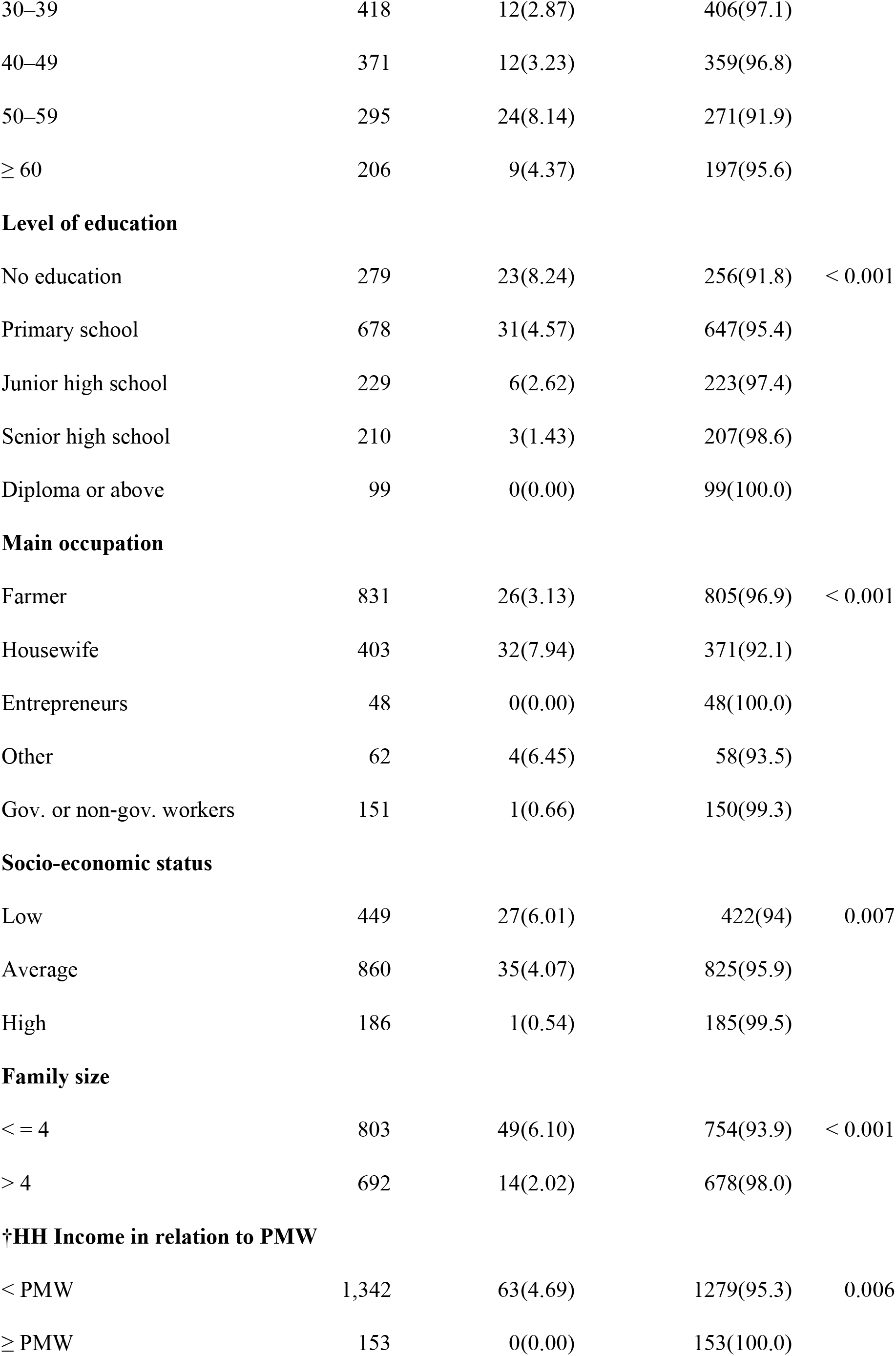
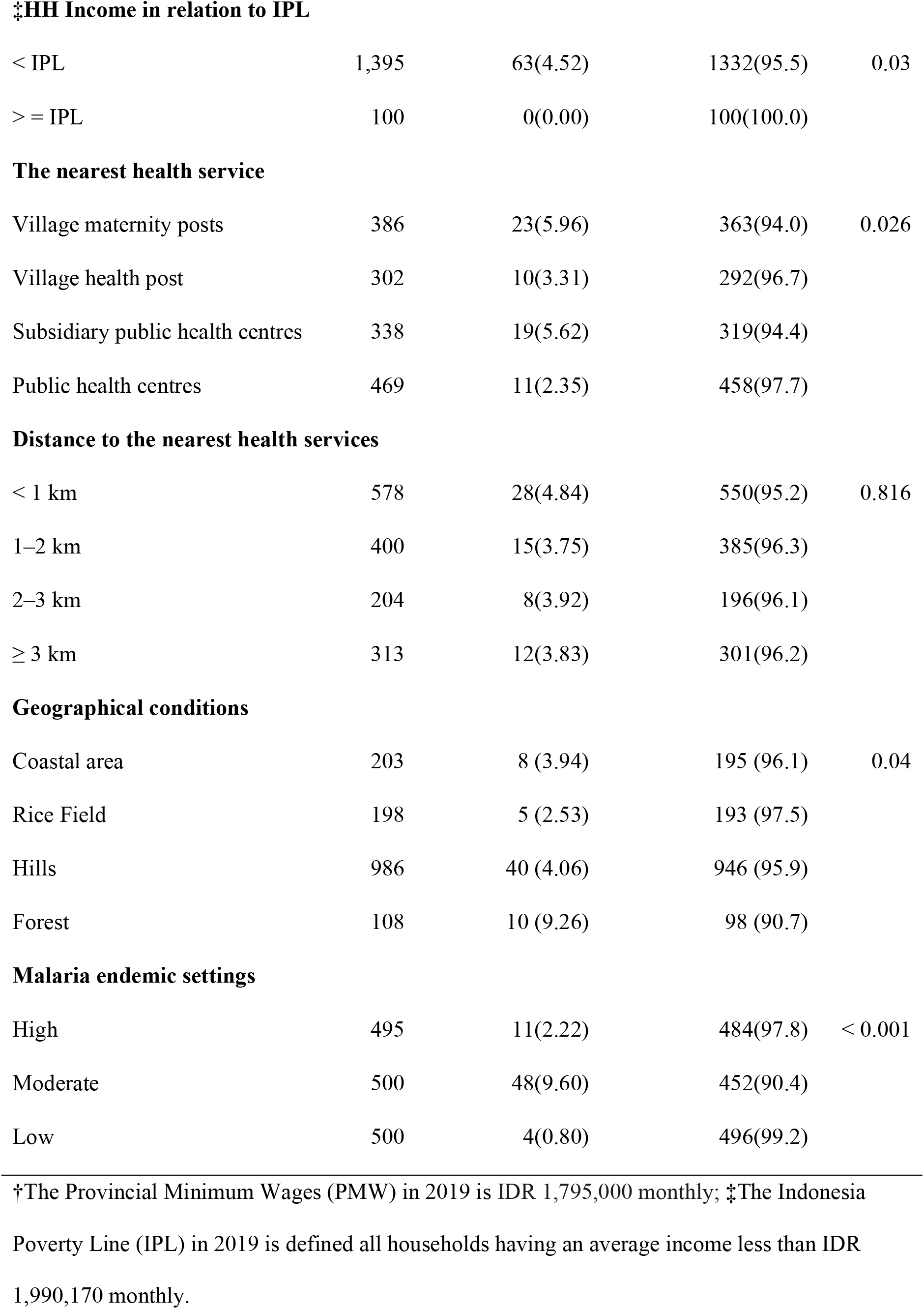
The ownership of mosquito nets by Socio-demographic and Environmental Characteristics of Respondents in East Nusa Tenggara Province (N = 1,495)

| Characteristics | Total | Ownership of mosquito nets, (n %) |  | p-value |
| --- | --- | --- | --- | --- |
|  |  | No | Yes |  |
| **Overall** | 1,495 | 63(4.21) | 1432(95.8) |  |
| **Gender** |  |  |  |  |
| Male | 727 | 27(3.71) | 700(96.3) | 0.349 |
| Female | 768 | 36(4.69) | 732(95.3) |  |
| **Age group** |  |  |  |  |
| < 30 | 205 | 6(2.93) | 199(97.1) | 0.005 |
| 30–39 | 418 | 12(2.87) | 406(97.1) |  |
| 40–49 | 371 | 12(3.23) | 359(96.8) |  |
| 50–59 | 295 | 24(8.14) | 271(91.9) |  |
| ≥ 60 | 206 | 9(4.37) | 197(95.6) |  |
| **Level of education** |  |  |  |  |
| No education | 279 | 23(8.24) | 256(91.8) | < 0.001 |
| Primary school | 678 | 31(4.57) | 647(95.4) |  |
| Junior high school | 229 | 6(2.62) | 223(97.4) |  |
| Senior high school | 210 | 3(1.43) | 207(98.6) |  |
| Diploma or above | 99 | 0(0.00) | 99(100.0) |  |
| **Main occupation** |  |  |  |  |
| Farmer | 831 | 26(3.13) | 805(96.9) | < 0.001 |
| Housewife | 403 | 32(7.94) | 371(92.1) |  |
| Entrepreneurs | 48 | 0(0.00) | 48(100.0) |  |
| Other | 62 | 4(6.45) | 58(93.5) |  |
| Gov. or non-gov. workers | 151 | 1(0.66) | 150(99.3) |  |
| **Socio-economic status** |  |  |  |  |
| Low | 449 | 27(6.01) | 422(94) | 0.007 |
| Average | 860 | 35(4.07) | 825(95.9) |  |
| High | 186 | 1(0.54) | 185(99.5) |  |
| **Family size** |  |  |  |  |
| < = 4 | 803 | 49(6.10) | 754(93.9) | < 0.001 |
| > 4 | 692 | 14(2.02) | 678(98.0) |  |
| **†HH Income in relation to PMW** |  |  |  |  |
| < PMW | 1,342 | 63(4.69) | 1279(95.3) | 0.006 |
| ≥ PMW | 153 | 0(0.00) | 153(100.0) |  |
| ‡HH Income in relation to IPL |  |  |  |  |
| < IPL | 1,395 | 63(4.52) | 1332(95.5) | 0.03 |
| > = IPL | 100 | 0(0.00) | 100(100.0) |  |
| The nearest health service |  |  |  |  |
| Village maternity posts | 386 | 23(5.96) | 363(94.0) | 0.026 |
| Village health post | 302 | 10(3.31) | 292(96.7) |  |
| Subsidiary public health centres | 338 | 19(5.62) | 319(94.4) |  |
| Public health centres | 469 | 11(2.35) | 458(97.7) |  |
| Distance to the nearest health services |  |  |  |  |
| < 1 km | 578 | 28(4.84) | 550(95.2) | 0.816 |
| 1–2 km | 400 | 15(3.75) | 385(96.3) |  |
| 2–3 km | 204 | 8(3.92) | 196(96.1) |  |
| ≥ 3 km | 313 | 12(3.83) | 301(96.2) |  |
| Geographical conditions |  |  |  |  |
| Coastal area | 203 | 8 (3.94) | 195 (96.1) | 0.04 |
| Rice Field | 198 | 5 (2.53) | 193 (97.5) |  |
| Hills | 986 | 40 (4.06) | 946 (95.9) |  |
| Forest | 108 | 10 (9.26) | 98 (90.7) |  |
| Malaria endemic settings |  |  |  |  |
| High | 495 | 11(2.22) | 484(97.8) | < 0.001 |
| Moderate | 500 | 48(9.60) | 452(90.4) |  |
| Low | 500 | 4(0.80) | 496(99.2) |  |

†The Provincial Minimum Wages (PMW) in 2019 is IDR 1,795,000 monthly; ‡The Indonesia
Poverty Line (IPL) in 2019 is defined all households having an average income less than IDR 1,990,170 monthly.

The ownership and the use of mosquito nets in rural adults of ENTP is showed in Table 2. Overall, the percentage of participants with the possession of mosquito nets was high at 95.8% with a 95% confidence interval (CI): 94.8 – 96.8. The proportion of participants having LLINs was high, accounting for 71.6% with a 95% CI: 68.9 – 74.3, whilst the ownership of non-LLINs was only 42.7% with a 95% CI: 38.8 – 46.5. The use of any mosquito nets by rural adults was high, accounting for 82.3% with a 95% CI :80.2-84.5, however only about half of participants (54.8% with a 95% CI: 51.4 – 58.2) utilised LLINs. This was worsened for non-LLINs. Merely about the quarter of participants (27.6% with a 95% CI: 23.2 – 31.9) applied non-LLINs for preventing malaria.

**Table 2.** The ownership and the use of mosquito nets of the respondents in East Nusa Tenggara Province (N = 1,495)

| # | Questions | n | % (95%CI) |
| --- | --- | --- | --- |
| 1 | The ownership of mosquito nets |  |  |
|  | Yes | 1432 | 95.8(94.8, 96.8) |
|  | No | 63 | 4.20(0.00, 9.20) |
| 2 | Types of mosquito nets |  |  |
|  | Only LLINs | 794 | 53.1(49.6, 56.6) |
|  | Only Non LLINs | 362 | 24.2(19.8, 28.6) |
|  | LLINs and Non-LLINs | 276 | 18.5(13.9, 23.1) |
| 3 | Respondent sleeping under bed net last night |  |  |
|  | Yes | 1231 | 82.3(80.2, 84.4) |
|  | No | 264 | 17.6(13.0, 22.2) |
| 4 | Types of bed net used last night |  |  |
|  | LLINs | 819 | 54.8(51.4, 58.2) |
|  | Non-LLINs | 412 | 27.6(23.3, 31.9) |
|  | Respondents have owned any mosquito nets | 1432 | 95.8(94.7, 96.8) |
|  | Respondents have owned non-LLINs | 638 | 42.7(38.8, 46.5) |
|  | Respondents have owned LLINs | 1070 | 71.6(68.9, 74.3) |
|  | Respondents sleeping under Non LLINs last night | 412 | 27.6(23.2, 31.9) |
|  | Respondents sleeping under LLINs last night | 819 | 54.8(51.4, 58.2) |
|  | Respondent sleeping under Any bed net last night | 1231 | 82.3(80.2, 84.5) |

Factors associated with the use of any mosquito nets in rural of ENTP was presented in Table 3. After controlling all confounding variables in the multivariable analysis, the following results were obtained. Having at least diploma education level (adjusted odds ratio (AOR) = 5.68, 95% CI: 2.6 – 12.41), senior high school education level (AOR = 5.41, 95% CI: 3.07– 9.53), junior high school education level (AOR= 2.87, 95% CI: 1.78 – 4.63), primary education level (AOR = 2.52, 95% CI: 1.79 – 3.55), living in low endemic settings (AOR = 10.1, 95% CI: 6.17 – 16.5), living in moderate endemic settings (AOR = 2.09, 95% CI:1.50 -2.90), having small family members (AOR = 1.73, 95% CI: 1.30 - 2.30), living closed to rice field (AOR= 2.36, 95% CI: 1.30 - 4.30), living closed to hills (AOR= 2.10, 95% CI: 1.32 – 3.36) were considerably associated with the use of any mosquito nets.

**Table 3.** Factors associated with the use of any mosquito nets in East t Nusa Tenggara Province (N = 1,495)

| Characteristics | Using of any mosquito nets |  |  |  |
| --- | --- | --- | --- | --- |
|  | No at risk | n (%) | COR<sup>†</sup> | AOR<sup>‡</sup> |
| **Overall** | 1,495 | 1231 (82.3) |  |  |
| **Gender** |  |  |  |  |
| Male | 727 | 606 (83.4) | 1.15 (0.88, 1.50) | 1.19 (0.89, 1.59) |
| Female | 768 | 625 (81.4) | 1.00 | 1.00 |
| **Age group** |  |  |  |  |
| < 30 | 205 | 171 (83.4) | 1.37 (0.83, 2.24) | 0.84 (0.48, 1.45) |
| 30–39 | 418 | 359 (85.9) | 1.65 (1.07, 2.55) | 1.32 (0.82, 2.12) |
| 40–49 | 371 | 298 (80.3) | 1.11 (0.73, 1.69) | 0.97 (0.61, 1.52) |
| 50–59 | 295 | 241 (81.7) | 1.21 (0.78, 1.89) | 1.05 (0.66, 1.68) |
| ≥ 60 | 206 | 162 (78.6) | 1.00 | 1.00 |
| **Level of education** |  |  |  |  |
| No education | 279 | 189 (67.7) | 1.00 | 1.00 |
| Primary school | 678 | 570 (84.1) | 2.51 (1.82, 3.48) | 2.52 (1.79, 3.55) |
| Junior high school | 229 | 195 (85.2) | 2.73 (1.76, 4.25) | 2.87 (1.78, 4.63) |
| Senior high school | 210 | 188 (89.5) | 4.07 (2.45, 6.76) | 5.41 (3.07, 9.53) |
| Diploma or above | 99 | 89 (89.9) | 4.24 (2.10, 8.54) | 5.68 (2.6, 12.41) |
| **Socio-economic status** |  |  |  |  |
| Low | 449 | 373 (83.1) | 1.52 (1.00, 2.31) | 2.18 (1.34, 3.54) |
| Average | 860 | 716 (83.3) | 1.54 (1.05, 2.26) | 2.14 (1.39, 3.28) |
| High | 186 | 142 (76.3) | 1.00 | 1.00 |
| **Family size** |  |  |  |  |
| < = 4 | 803 | 691 (86.1) | 1.74 (1.33, 2.27) | 1.73 (1.30, 2.30) |
| > 4 | 692 | 540 (78.0) | 1.00 | 1.00 |
| **Geographical conditions** |  |  |  |  |
| Coastal area | 203 | 146 (71.9) | 1.18 (0.71, 1.96) | 0.84 (0.48, 1.45) |
| Rice Field | 198 | 171 (86.4) | 2.91 (1.64, 5.17) | 2.36 (1.30, 4.30) |
| Hills | 986 | 840 (85.2) | 2.64 (1.70, 4.12) | 2.10 (1.32, 3.36) |
| Forest | 108 | 74 (68.5) | 1.00 | 1.00 |
| **Malaria endemic settings** |  |  |  |  |
| High | 495 | 334 (67.5) | 1.00 | 1.00 |
| Moderate | 500 | 418 (83.6) | 2.46 (1.82, 3.33) | 2.09 (1.50, 2.90) |
| Low | 500 | 479 (95.8) | 11.0 (6.83, 17.7) | 10.1 (6.17, 16.5) |

<sup>†</sup>COR = crude odds ratio, <sup>‡</sup>AOR = adjusted odds ratio; Main occupation, household income, the nearest health facilities, and the distance to the nearest health facilities were not significant in the final model.

Factors associated with the use of non-LLINs in rural of ENTP was depicted in Table 4. After controlling all confounding variables in multivariable analysis, the following results were obtained. Having at least diploma education (adjusted odds ratio (AOR) = 2.37, 95% CI: 1.14 – 4.93), senior high school education level (AOR = 2.58, 95% CI: 1.57– 4.23), junior high school education level (AOR = 2.69, 95% CI: 1.69 – 4.30), primary education level (AOR = 2.26, 95% CI: 1.52 – 3.35), living in low endemic settings (AOR = 11.1, 95% CI: 7.32 – 16.7), living in moderate endemic settings (AOR = 3.52, 95% CI:2.27 -5.46), living closed to public health centre (AOR = 2.25, 95% CI: 1.58 – 3.20), living closed to subsidiary public health centre (AOR = 1.79, 95% CI: 1.22 – 2.63), living closed to village health post (AOR = 2.06, 95% CI: 1.39 – 3.05), living less than 1 km from the nearest health service (AOR = 1.51, 95% CI: 1.05 – 2.18), living between 1 to 2 km from the nearest health service (AOR = 2.43, 95% CI: 1.67 – 3.54), living between 2 to 3 km from the nearest health service (AOR = 2.16, 95% CI: 1.42 – 3.30), were significantly associated with the use of non-LLINs.

**Table 4.** Factors associated with the use of non-LLINs in East t Nusa Tenggara Province (N = 1495)

| Characteristics | Using of non - LLINs |  |  |  |
| --- | --- | --- | --- | --- |
|  | No at risk | n (%) | COR<sup>†</sup> | AOR<sup>‡</sup> |
| **Overall** | 1,495 | 412 (27.6) |  |  |
| **Gender** |  |  |  |  |
| Male | 727 | 209 (28.7) | 1.12 (0.90, 1.41) | 0.96 (0.70, 1.31) |
| Female | 768 | 203 (26.4) | 1.00 | 1.00 |
| **Age group** |  |  |  |  |
| < 30 | 205 | 57 (27.8) | 1.34 (0.86, 2.10) | 1.02 (0.61, 1.68) |
| 30–39 | 418 | 133 (31.8) | 1.62 (1.10, 2.39) | 1.17 (0.77, 1.79) |
| 40–49 | 371 | 90 (24.3) | 1.11 (0.74, 1.67) | 0.91 (0.59, 1.41) |
| 50–59 | 295 | 86 (29.2) | 1.43 (0.95, 2.16) | 1.18 (0.76, 1.83) |
| ≥ 60 | 206 | 46 (22.3) | 1.00 | 1.00 |
| **Level of education** |  |  |  |  |
| No education | 279 | 39 (14.0) | 1.00 | 1.00 |
| Primary school | 678 | 188 (27.7) | 2.36 (1.62, 3.45) | 2.26 (1.52, 3.35) |
| Junior high school | 229 | 73 (31.9) | 2.88 (1.86, 4.46) | 2.69 (1.69, 4.30) |
| Senior high school | 210 | 70 (33.3) | 3.08 (1.97, 4.80) | 2.58 (1.57, 4.23) |
| Diploma or above | 99 | 42 (42.4) | 4.53 (2.69, 7.65) | 2.37 (1.14, 4.93) |
| **Main occupation** |  |  |  |  |
| Farmer | 831 | 214 (25.8) | 1.00 | 1.00 |
| Housewife | 403 | 105 (26.1) | 1.02 (0.77, 1.33) | 0.96 (0.67, 1.38) |
| Entrepreneurs | 48 | 10 (20.8) | 0.76 (0.37, 1.55) | 0.78 (0.37, 1.65) |
| Other | 62 | 14 (22.6) | 0.84 (0.46, 1.56) | 0.93 (0.48, 1.81) |
| Gov. or non-gov. workers | 151 | 69 (45.7) | 2.43 (1.70, 3.46) | 1.81 (1.06, 3.11) |
| **The nearest health service** |  |  |  |  |
| Village maternity posts | 386 | 66 (17.1) | 1.00 | 1.00 |
| Village health post | 302 | 86 (28.5) | 1.93 (1.34, 2.78) | 2.06 (1.39, 3.05) |
| Subsidiary public health centres | 338 | 88 (26.0) | 1.71 (1.19, 2.45) | 1.79 (1.22, 2.63) |
| Public health centres | 469 | 172 (36.7) | 2.81 (2.03, 3.89) | 2.25 (1.58, 3.20) |
| **Distance to the nearest health services** |  |  |  |  |
| < 1 km | 578 | 140 (24.2) | 1.35 (0.96, 1.89) | 1.51 (1.05, 2.18) |
| 1–2 km | 400 | 140 (35.0) | 2.27 (1.60, 3.22) | 2.43 (1.67, 3.54) |
| 2–3 km | 204 | 72 (35.3) | 2.30 (1.54, 3.44) | 2.16 (1.42, 3.30) |
| ≥ 3 km | 313 | 60 (19.2) | 1.00 | 1.00 |
| **Geographical conditions** |  |  |  |  |
| Coastal area | 203 | 50 (24.6) | 1.28 (0.73, 2.25) | 0.56 (0.30, 1.04) |
| Rice Field | 198 | 82 (41.4) | 2.76 (1.60, 4.78) | 0.54 (0.28, 1.04) |
| Hills | 986 | 258 (26.2) | 1.39 (0.85, 2.26) | 0.37 (0.21, 0.65) |
| Forest | 108 | 22 (20.4) | 1.00 | 1.00 |
| **Malaria endemic settings** |  |  |  |  |
| High | 495 | 44 (8.89) | 1.00 | 1.00 |
| Moderate | 500 | 120 (24.0) | 3.24 (2.23, 4.69) | 3.52 (2.27, 5.46) |
| Low | 500 | 248 (49.6) | 10.1 (7.07, 14.4) | 11.1 (7.32, 16.7) |

<sup>†</sup>COR = crude odds ratio, <sup>‡</sup>AOR = adjusted odds ratio; Main occupation, Social economic status, Household income, the nearest health facilities, and the distance to the nearest health facilities were not significant in the final model.

Factors associated with the use of LLINs in rural of ENTP was portrayed in Table 5. After controlling all confounding variables in multivariable analysis, the following results were obtained. Living in low endemic settings (AOR = 0.60, 95% CI:0.45 -0.80) and living closed to hills (AOR = 2.11, 95% CI: 1.37 – 3.25) were substantially associated with the use of LLINs.

**Table 5.** Factors associated with the use of LLINs in East t Nusa Tenggara Province (N = 1495)

| Characteristics | Using of LLINs |  |  |  |
| --- | --- | --- | --- | --- |
|  | No at risk | n (%) | COR<sup>†</sup> | AOR<sup>‡</sup> |
| **Overall** | 1,495 | 819 (54.8) |  |  |
| **Gender** |  |  |  |  |
| Male | 727 | 397 (54.6) | 0.99 (0.81, 1.21) | 1.1 (0.89, 1.37) |
| Female | 768 | 422 (54.9) | 1.00 | 1.00 |
| **Age group** |  |  |  |  |
| < 30 | 205 | 114 (55.6) | 0.97 (0.66, 1.44) | 1.00 (0.66, 1.50) |
| 30–39 | 418 | 226 (54.1) | 0.91 (0.65, 1.28) | 1.01 (0.71, 1.44) |
| 40–49 | 371 | 208 (56.1) | 0.99 (0.70, 1.40) | 1.02 (0.72, 1.45) |
| 50–59 | 295 | 155 (52.5) | 0.86 (0.60, 1.23) | 0.90 (0.62, 1.30) |
| ≥ 60 | 206 | 116 (56.3) | 1.00 | 1.00 |
| **Geographical conditions** |  |  |  |  |
| Coastal area | 203 | 96 (47.3) | 0.97 (0.61, 1.54) | 1.14 (0.70, 1.86) |
| Rice Field | 198 | 89 (44.9) | 0.88 (0.55, 1.41) | 1.51 (0.91, 2.52) |
| Hills | 986 | 582 (59) | 1.55 (1.04, 2.31) | 2.11 (1.37, 3.25) |
| Forest | 108 | 52 (48.1) | 1.00 | 1.00 |
| **Malaria endemic settings** |  |  |  |  |
| High | 495 | 290 (58.6) | 1.00 | 1.00 |
| Moderate | 500 | 298 (59.6) | 1.04 (0.81, 1.34) | 0.98 (0.74, 1.31) |
| Low | 500 | 231 (46.2) | 0.61 (0.47, 0.78) | 0.60 (0.45, 0.80) |

<sup>†</sup>COR = crude odds ratio, <sup>‡</sup>AOR = adjusted odds ratio; Education level, Main occupation, Social economic status, Household income, family size, the nearest health facilities, and the distance to the nearest health facilities were not significant in the final model.

## DISCUSSION

To our knowledge this is the first population-based study to explore the ownership and the utilisation of mosquito nets in rural adult of East Nusa Tenggara Province Indonesia. This study shows that the ownership of any mosquito nets was high in this area, however the utilisation of this nets particularly long-lasting insecticide treated net (LLINs) was low. This study also demonstrated that the high coverage of LLINs was not followed by the high use of LLINs. The prominent factors associated with a higher likelihood to utilise mosquito nets were low malaria endemic settings, higher education level, and better socio-economic status.

The difference in coverage of LLINs and the use of LLINs was consistent with previous findings in some other countries ^21–24^. Our study shows that 72% percent of rural population have the possession of LLINs. This coverage was just below the coverage that recommended by WHO. Improving the coverage of these treated nets is critical to boost malaria elimination in this setting. This study also further demonstrated that the use of LLINs by rural population was low. The improvement of awareness of rural community to use LLINs is critical considering that the treated nets have been applied by the Indonesian government as the main intervention to control malaria in this country ^25^. Failure to educate community on the benefit of the treated nets led to misuse of them including for food storage as indicated in other research ^26^.

The results of this study showed that the use of mosquito nets was significantly higher in low MES compared to high MES. This result is consistent with findings in other settings ^23,27,28^. The high proportion of adults’ people in low MES sleeping under mosquito nets might be contributed to high awareness of malaria for those communities. Previous studies showed that adults in low MES in this region has high malaria awareness compared to those in high MES ^13^. The finding of this study is unexpected by the authors since the national government of Indonesia has allocated more resources in distributing mosquito net for high MES in the Eastern part of Indonesia including in this area since 2005 until now ^5,25^. This signalled that the distribution of mosquito nets in this area might not be followed by the adequate health promotion activities to encourage local community taking the benefit of the mosquito net in preventing malaria.

The present study demonstrated that the use of any mosquito nets was in line with the increase of education level of participants. This is consistent with other studies ^29–32^, indicating that there was a positive association between education level and the use of mosquito nets for preventing malaria. This study revealed that the likelihood of the use of mosquito net for participants having at least diploma education level was almost six times higher than those without education level. Participants having senior high school education level were five times higher to use mosquito net compared with those having no education level. The possible explanation for this could be that people with higher education level have a high level of malaria knowledge ^16,33^, and they have many opportunities to be exposed to multiple sources of information ^34^, so that they could understand the benefit of sleeping under mosquito net for preventing malaria. This research has demonstrated that more than half of rural adults in this province have low level of education (no formal school or only primary school) and other studies indicated that children dropping out of school was high in this area ^35,36^. These situations need to be addressed by local authority to encourage rural population using mosquito nets to support malaria elimination effort in this area.

One of the important aspects to assess the benefit of the health intervention programs is the socio-economic status of the community. The intervention programs should reach the poor as many times as they do to the relatively rich people. In this research, we found that the ownership of mosquito nets rose with the increasing of socio-economic status of participants. In contrast to the possession of mosquito nets, this study demonstrated that the poorest was more likely to use mosquito nets compared to the richest group. This finding is consistent with other finding in other settings ^37,38^. The high use of mosquito nets in the poor group might be connected with their perceived vulnerability as perceived susceptibility to malaria has been revealed to be higher in poor families. Moreover, people with high SES often have easy access to other methods for preventing malaria and might therefore not use of mosquito nets. One study indicated that the richest group living in houses with have screening in door and window often believed that they were adequately shielded from mosquito bites and therefore they did not use mosquito nets ^39^.

While this study provides some significant findings to guide malaria elimination programs in this province, it also has some weaknesses. First, the ownership and the utilisation of mosquito nets were assessed based on the information provided by one adult in a household. Each participant was asked how many mosquito nets they have and whether all family members slept under mosquito nets in the night prior survey. The response to these questions might prone to recall bias, which is a common type of bias in a cross-sectional study design. Secondly, this study was conducted during the rain season in this area. Similarly, the other provinces in Indonesia, this province has two seasons, rain and dry season. The behaviour of rural population might show a different pattern in the use of mosquito nets, as indicated in other studies ^40^. Therefore, the comprehensive pattern in the use mosquito nets throughout the year could not be revealed by this study.

In conclusion, this study demonstrated that the high coverage of mosquito nets particularly LLINs is not followed by the high use of this treated net. The important factors affecting the utilisation of mosquito nets were malaria endemic settings and education level of participants. To boost malaria elimination in ENTP, the coverage of any mosquito nets should be high. Health education promotion to improve the awareness rural community of the using of these nets should be prioritized for those living in high MES and low education level.

## Data Availability

All data produced in the present study are available upon reasonable request to the authors

## Acknowledgments

We would like to thank the Australia Awards Scholarship for supporting this research and School of Health Sciences, Swinburne University of Technology for providing funding for primary data collection for this study. The funders had no role in the designing of the study, data collection, analysis, or interpretation of data, or writing the paper. We further would like to express our gratitude to the Health Ministry of Indonesia, the governor of ENTP, head of East Sumba, Belu, and East Manggarai district, nine head of sub-districts, and forty-nine village leaders for allowing this research to be conducted in their regions.

## Author Contributions

Conceptualization: RDG. Data curation: RDG, FMAI. Formal analysis: RDG, FMAI. Funding acquisition: RDG, FMAI. Methodology: RDG. Project administration: RDG, ML. Writing - original draft: RDG. Writing - review & editing: RDG, ML, FMAI

---

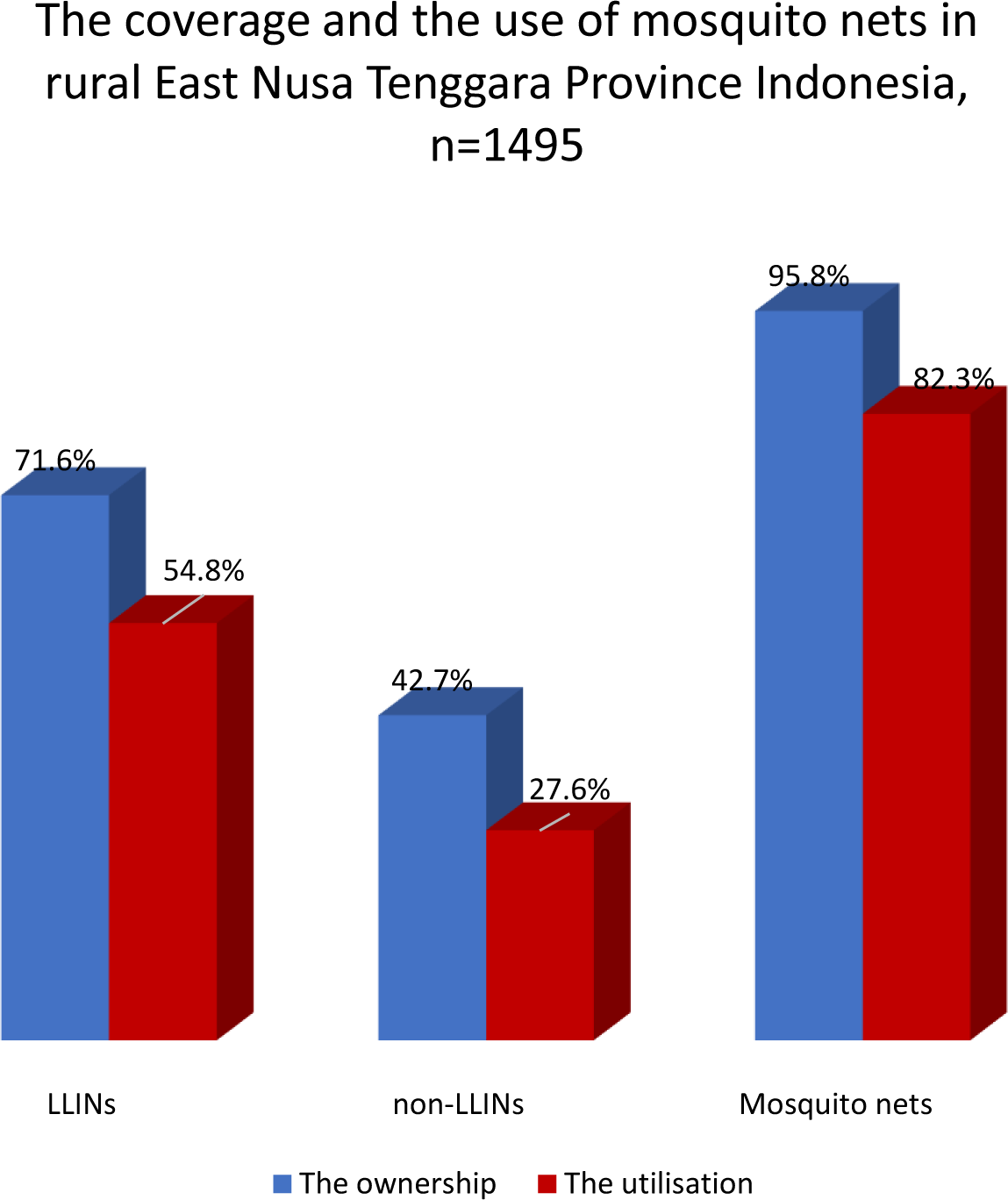

